# REPERCUSSIONS OF THE COVID-19 PANDEMIC ON THE MENTAL HEALTH OF PREGNANT AND PUERPERAL WOMEN: A SYSTEMATIC REVIEW

**DOI:** 10.1101/2020.08.17.20176560

**Authors:** Leticia Genova Vieira, Emerson Lucas Junior Silva Camargo, Guilherme Schneider, Gabrielly Pereira Rocatti da Silva, Micaella Thomazini, Matheus Arantes Possani, Matheus Rozário Matioli, Aline Raquel de Sousa Ibiapina

## Abstract

**Background:** The *Coronavirus Disease 2019* (COVID-19) pandemic has caused negative impacts on the physical and mental health of the population worldwide. Pregnant and puerperal women comprise the population most vulnerable to impacts on mental health.

**Objective:** To synthesize the scientific evidence on the repercussions of the COVID-19 pandemic on the mental health of pregnant and puerperal women.

**Methods:** Systematic review focused on answering the question “what is the impact of the COVID-19 pandemic on the mental health of pregnan and puerperal women?”. In order to perform the search of the studies, we used combinations among the keywords: *pregnan*, puerper*, prenatal, perinatal, “mental health”, COVID-19, SARS-CoV-2*. In total, we identified 150 studies from the databases and 14 studies were selected from preprints. We identified another four studies through manual search, totaling 18 studies to compose the final sample of this review.

**Results:** Anxiety and depression were the main outcomes found, being shown in 15 and 11 studies, respectively. Other outcomes found in more than one study were: concerns related to several factors, loneliness, stress and fear.

**Conclusion:** From this review, we can infer that the COVID-19 pandemic has impacted the mental health of pregnant and puerperal women, with depression and anxiety being the most frequent changes. The social detachment, the media pressure, the fear of contracting the infection, the economic scenario and the rupture of family rituals are shown as intensifying factors of psychological distress, thus causing changes in the mental health of these women.

## 1. INTRODUCTION

The onset of emerging infectious diseases has been considered a major public health problem worldwide. Atypical cases of pneumonia, severe respiratory disease and sudden impairment of other organs appeared in the city of Wuhan, Hubei province, China, in December 2019, caused by Severe Acute Respiratory Syndrome Coronavirus 2, SARS-CoV-2, etiologic agent of Coronavirus Disease 2019, COVID-19 [1].

With the exponential growth of COVID-19 on a transcontinental basis, the World Health Organization (WHO) declared, on March 11^th^, 2020, that COVID-19 qualified as a pandemic. Although the disease has low lethality, which ranges from 0.2% to 14.8%, its worsening is associated with an increase in the age group and the clinical impairment of pre-existing diseases. Its transmissibility is high, reaching about 80% of mild cases of acute respiratory syndrome, where between 5% and 10% of cases progress to severe symptoms of respiratory failure [2]. Respiratory secretions are the main source of the spread of COVID-19 [3]. Moreover, COVID-19 is capable of causing neurological, hepatic, respiratory and enteric changes [4].

Since the WHO declaration, health agencies have adopted measures to protect and prevent the spread of the illness, such as social detachment, the use of personal protective equipment (PPE) and non-pharmaceutical strategies, such as hand washing [5]. The adoption of prevention and control measures are part of the strategic actions aimed at the population or groups with a higher risk of infection, and the adoption of measures to halt the spread of the virus has caused an impact on health results, mainly on the mental health of the population, especially in pregnant and puerperal women [6].

Women in gestational and puerperal periods are groups considered to be at risk, given the greater chances for serious complications and lethality [7]. Accordingly, the primary and hospital health care services adopted the reorganization of care flows and specific strategies for the clinical-care management of these women, aiming at reducing the number of cases of COVID-19. In view of the above, the assistance during childbirth, birth and follow-up of the parturient woman must comply with all technical recommendations in symptomatic, asymptomatic or positive cases for COVID-19, aiming at protecting the health of women and newborns [8].

The rapid spread, the lack of discovery of effective treatment, the unpredictability of the duration of the pandemic, the disclosure of false information, known as *fake news*, and the shortage of understanding of the population in complying with the recommendations of health authorities are characterized as factors risk factors for the impact on mental health [9].

The increase in *fake news* published on social media has also increased stress, fear and anxiety about the disease, since the way some information is exposed has generated negative consequences on the mental health of the population, in some moments of this pandemic, especially in women during gestational and puerperal periods [10].

In view of the pandemic scenario, the disclosure of scientific knowledge worldwide has been happening in a skilful way about information concerning COVID-19. Nevertheless, studies associated with the impact of the pandemic on the mental health of pregnant and puerperal women are scarce, as the event is recent, but highlights negative factors. Thus, this study had the objective of synthesizing the scientific evidence on the repercussions of the COVID-19 pandemic on the mental health of pregnant and puerperal women.

## 2. METHOD

This is a systematic literature review [11] without registration of a protocol, with a framework based on the recommendations of *The Joanna Briggs Institute* (JBI), which consists of the systematic and orderly grouping of scientific evidence on a given topic, in order to allow the association between factors and development of a condition (health outcome), based on a strictly structured process, thus ensuring that the results achieved reliable and significant.

We performed six steps to develop this review, namely: 1) definition of the objective and construction of the guiding question; 2) definition of the search strategy; 3) definition of the inclusion and exclusion criteria and the data to be extracted from the studies; 4) selection of the studies; 5) characterization of the studies and synthesis of the pertinent data; 6) interpretation and presentation of the review.

The guiding question of this review “What is the impact of the COVID-19 pandemic on the mental health of pregnant and puerperal women?” was constructed with the aid of the PICOS strategy, as displayed in Table 1.

**Table 1.**
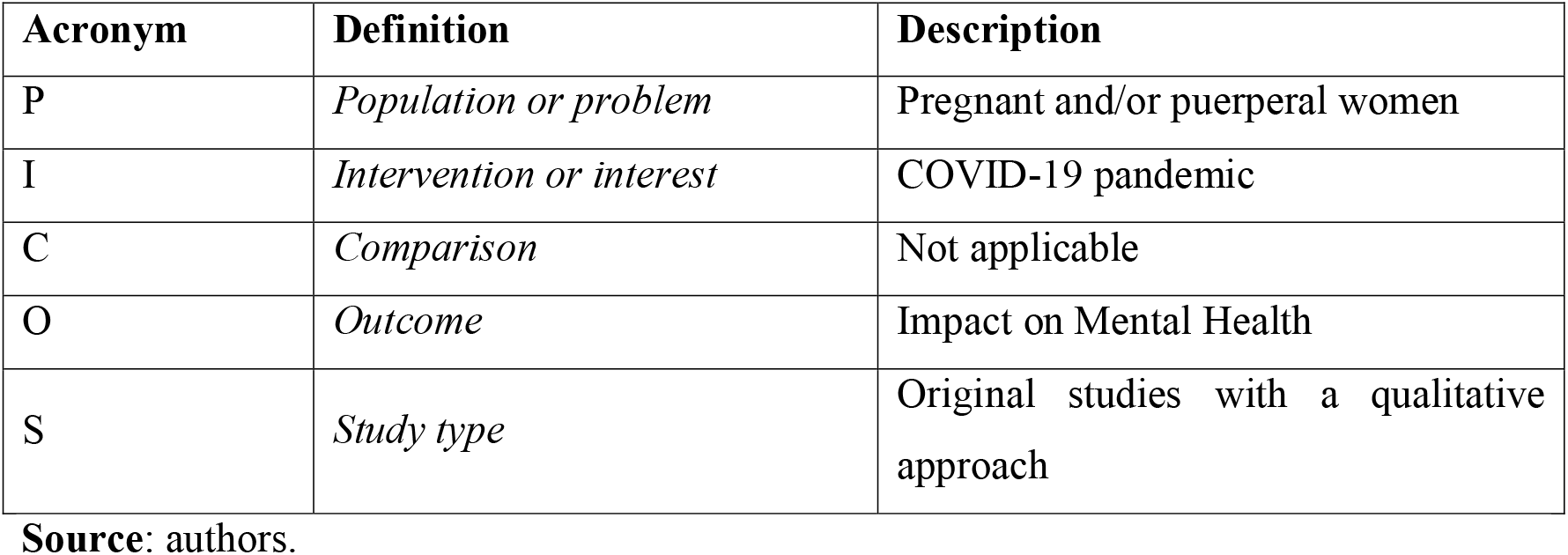
Application of the PICOS strategy for the construction of the research question. Ribeirão Preto, SP, Brazil, 2020.

| Acronym | Definition | Description |
| --- | --- | --- |
| P | *Population or problem* | Pregnant and/or puerperal women |
| I | *Intervention or interest* | COVID-19 pandemic |
| C | *Comparison* | Not applicable |
| O | *Outcome* | Impact on Mental Health |
| S | *Study type* | Original studies with a qualitative approach |

**Source:** authors.

The bibliographical survey was held during the month of July 2020, taking place in the following electronic databases: MEDLINE via PubMed portal of the US National Library of Medicine, Web of Science (WoS), Excerpta Medica Database (EMBASE), SCOPUS and APA PsycNet, in addition to the MedRxiv and PsyArXiv preprint databases. In order to complement the present study with the other evidence regarding the topic, we performed a manual search.

For the MEDLINE, WoS, EMBASE, SCOPUS, APA PsycNet databases and for the PsyArXiv preprint database, we used the following combination of keywords and Boolean operators: *(pregnan* OR puerper* OR prenatal OR perinatal)* AND *“mental health”AND (COVID-19 OR SARS-CoV-2)*. For the MedRxiv preprint database, we used the following search strategy: *(pregnancy OR pregnant OR puerperium OR puerperal OR prenatal OR perinatal) AND “mental health” AND (COVID-19 OR SARS-CoV-2)*.

We should underline that no filters were added as to the language and period of publication of scientific studies, with a view to encompassing the largest possible amount of evidence related to the problem in question.

Accordingly, we identified a total of 150 scientific works in the databases and preprint. Table 2 shows the number of studies retrieved from each queried database.

**Table 2.** Number of studies identified in each database regarding the impact of the COVID-19 pandemic on the mental health of pregnant and puerperal women. Ribeirão Preto, SP, Brazil, 2020.

| **Electronic databases and preprint** | **Number of identified studies** |
| --- | --- |
| MEDLINE | 28 |
| WOS | 11 |
| EMBASE | 52 |
| SCOPUS | 34 |
| APA PsycNet | 7 |
| PsyArXiv | 2 |
| medRxiv | 16 |
| **TOTAL** | **150** |

**Source:** authors.

Among the 150 identified studies, 69 were removed due to duplication in at least two databases. Thus, the titles and abstracts of the 81 screened studies were analyzed, by two reviewers, independently and blindly. For any disagreements between the researchers during the analytic process, we called a third researcher for resolution. After the considerations of the third reviewer, we performed a manual search based on the references of the secondary articles obtained in the search in the databases.

We established inclusion criteria for all primary studies that evaluated the mental health of pregnant and puerperal women during the COVID-19 pandemic. In turn, the adopted exclusion criteria were: secondary studies, expert opinion articles, experience reports and other scientific works that did not meet the scope of this review.

Thus, we selected 18 studies to compose the final sample of this systematic literature review, aiming at synthesizing the evidence on the impact of the COVID-19 pandemic on the mental health of pregnant and puerperal women. The studies were identified according to adaptations of the recommendations of the *Preferred Reporting Items for Systematic Reviews and Meta-Analyses* (PRISMA), as displayed in Flowchart 1 [12].

**Flowchart 1.**
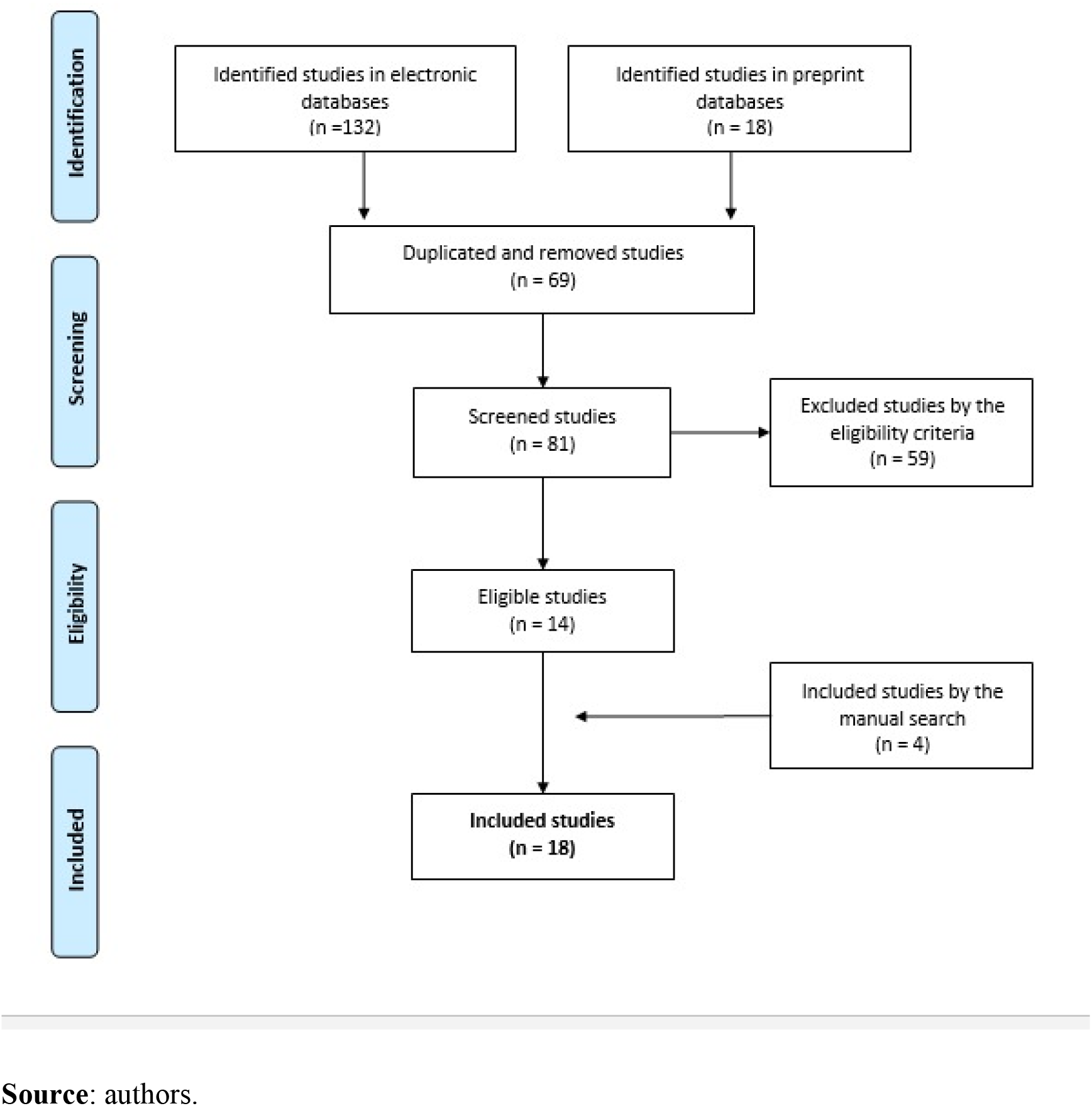
Schematized chart of the process of identification and selection of the studies to be included in this review, regarding the impact of the COVID-19 pandemic on the mental health of pregnant and puerperal women, according to the adaptation of the PRISMA model. Ribeirão Preto, SP, Brazil, 2020.

In this study, we analyzed the quality of the evidence levels of the included studies using the classification of the *Grading of Recommendations, Assessment, Development and Evaluation Working Group* (GRADE). The quality of the evidence obtained through the GRADE allows us to analyze the aggregated results, considering the design and the results of the included studies [13].

The data from the selected studies were extracted and grouped in a table (Table 3) containing specific information related to the research question, namely: author (s); year of publication; country where the research was held; component population and its main characteristics; study design; instruments and/or means used to evaluate mental health status; main outcomes related to the impact of the COVID-19 pandemic on the mental health of pregnant and puerperal women; level of evidence; and main limitations of the studies.

As this study used articles of public and free access, indexed in the electronic databases, the submission process in the Research Ethics Committee (CEP), according to the Resolution of the National Health Council (CNS) nº 466/2012, as well as the current ethical ethical standards, was not necessary.

## 3. RESULTS

We analyzed 18 studies that identified the impact on the mental health of pregnant and puerperal women during the period of the COVID-19 pandemic. In order to perform the qualitative synthesis of the selected articles, we drew up a table that summarizes the main characteristics of the included studies [14–31], containing information regarding the reference, the location and the population, the design, the outcome, the level of evidence and the limitations.

**Table 3.**
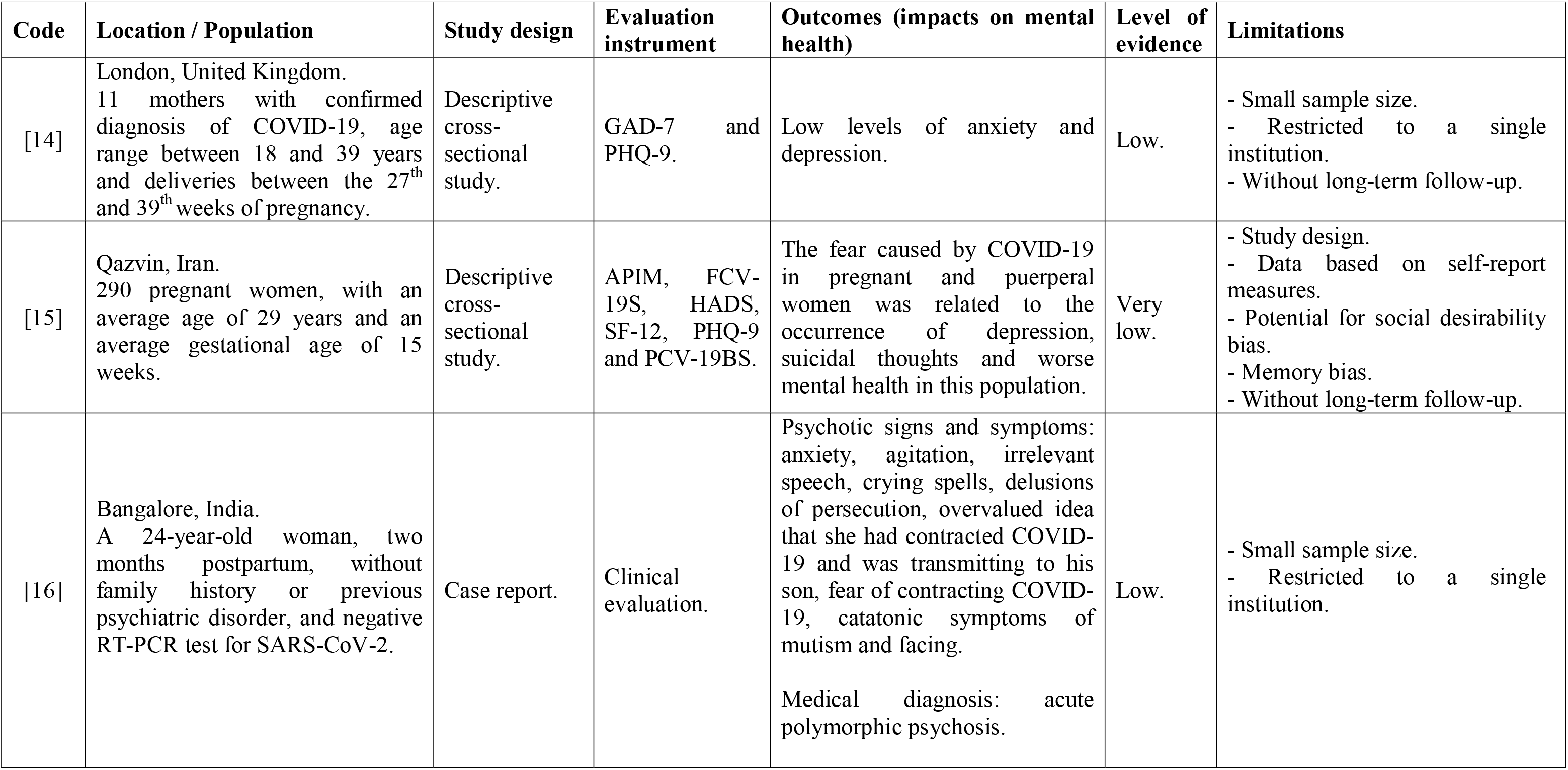
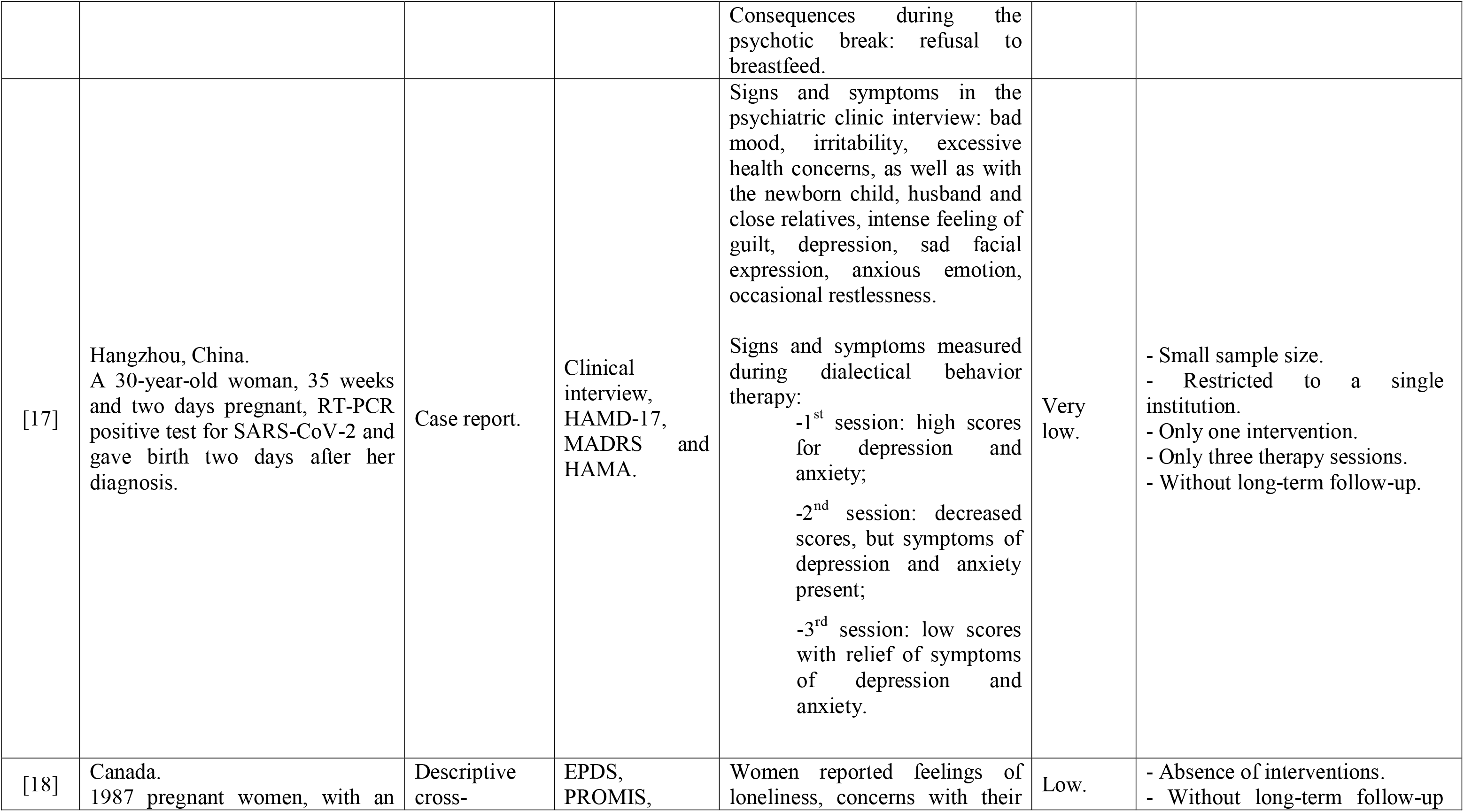
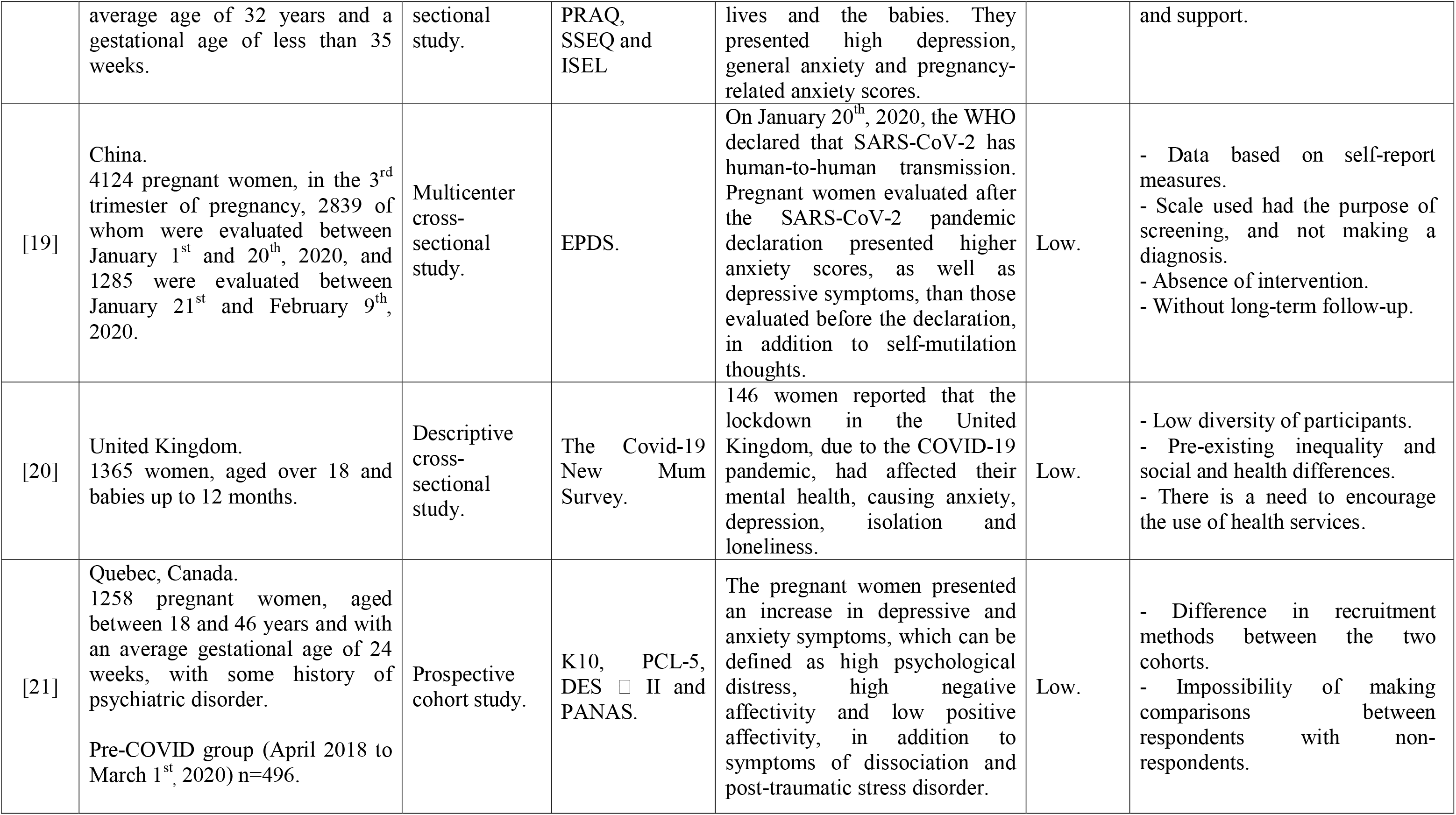
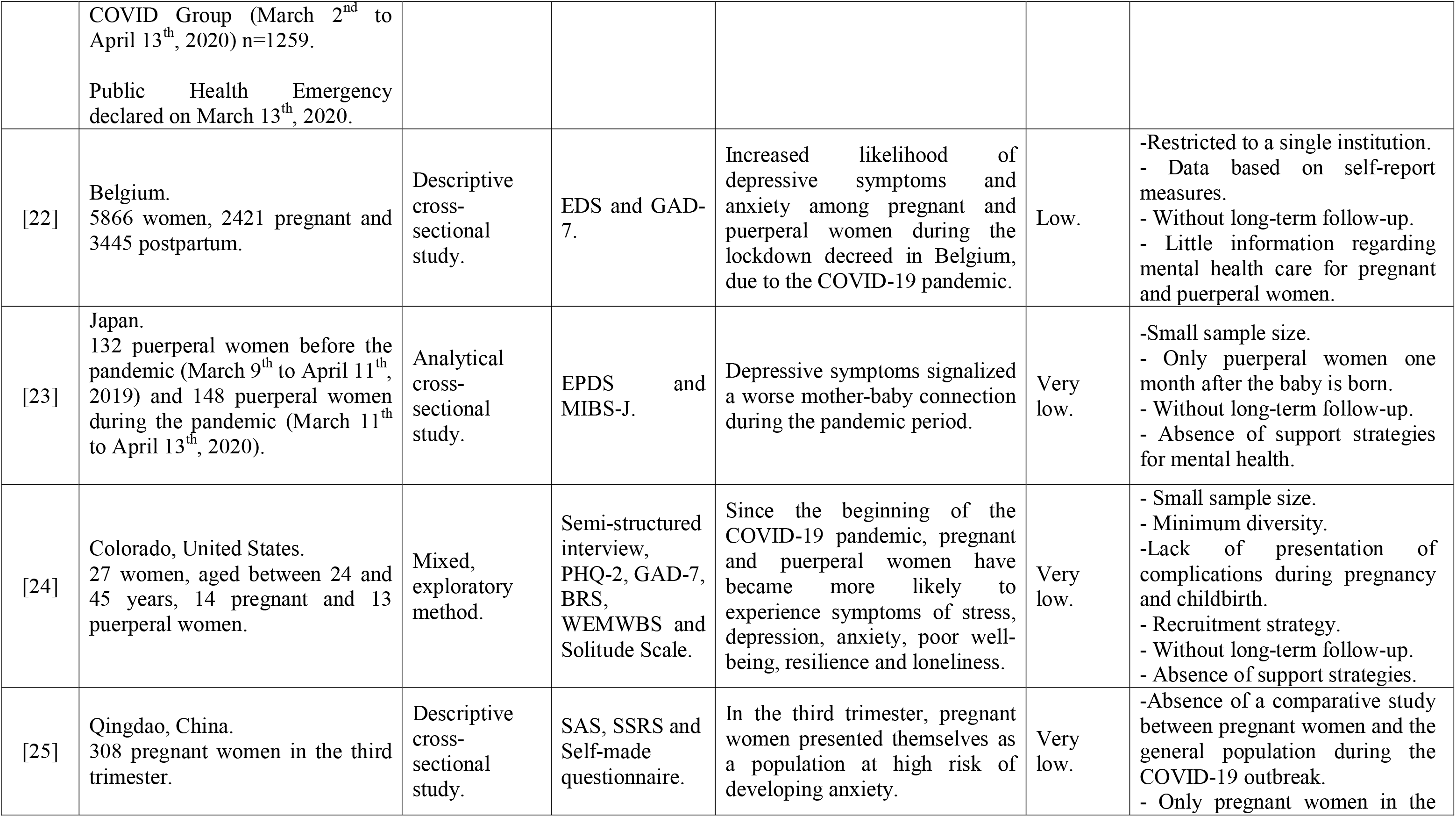
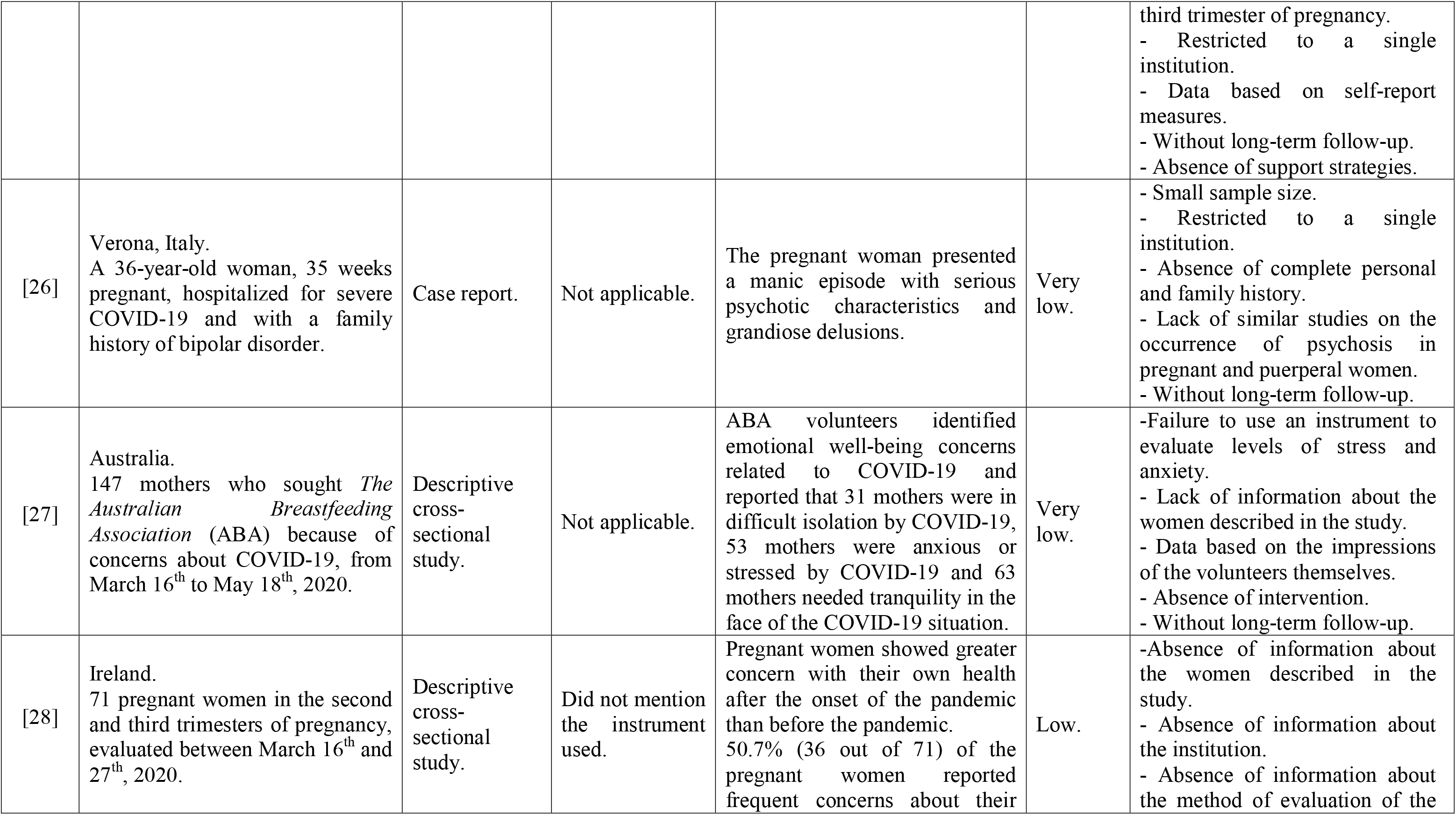
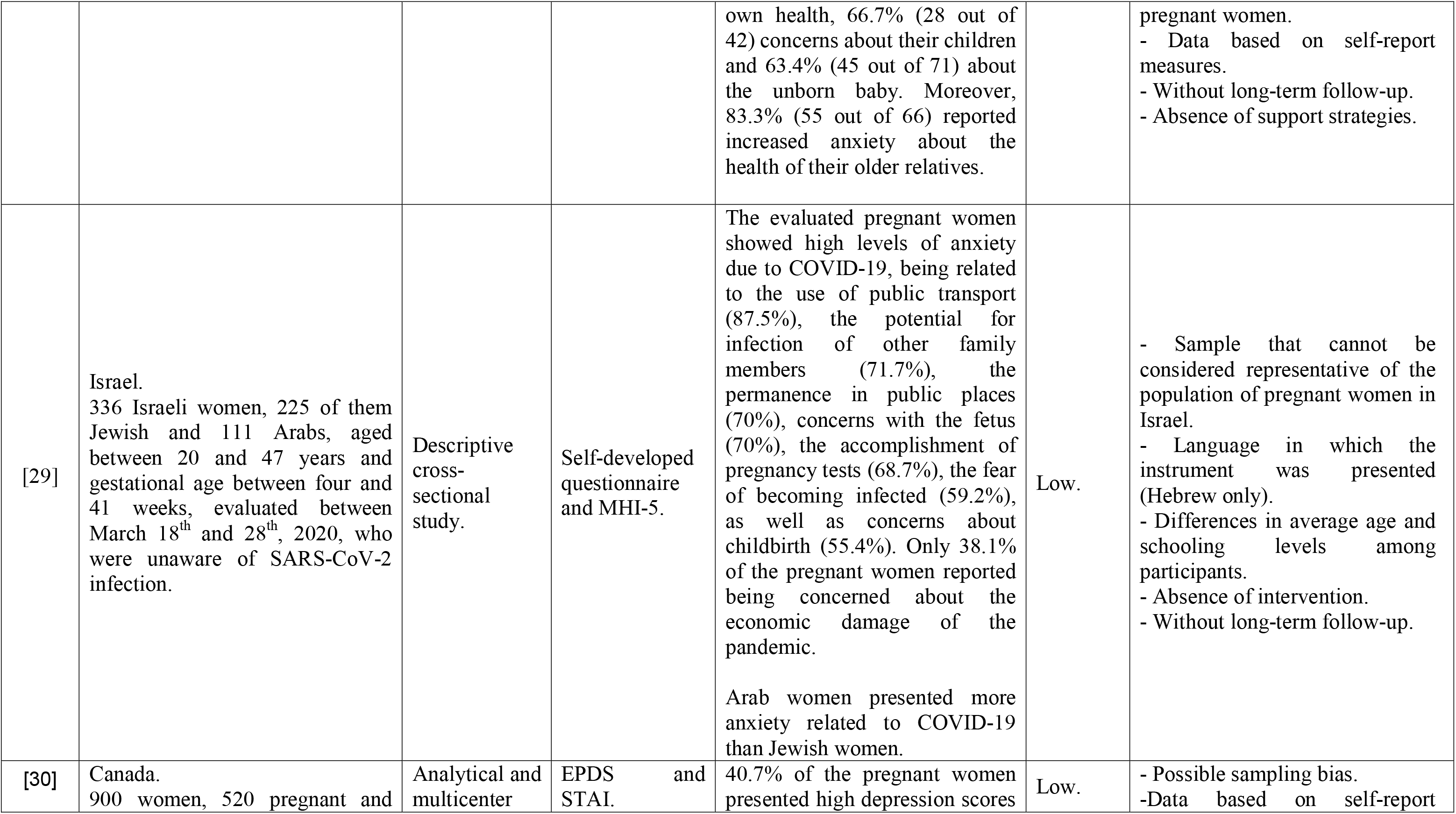
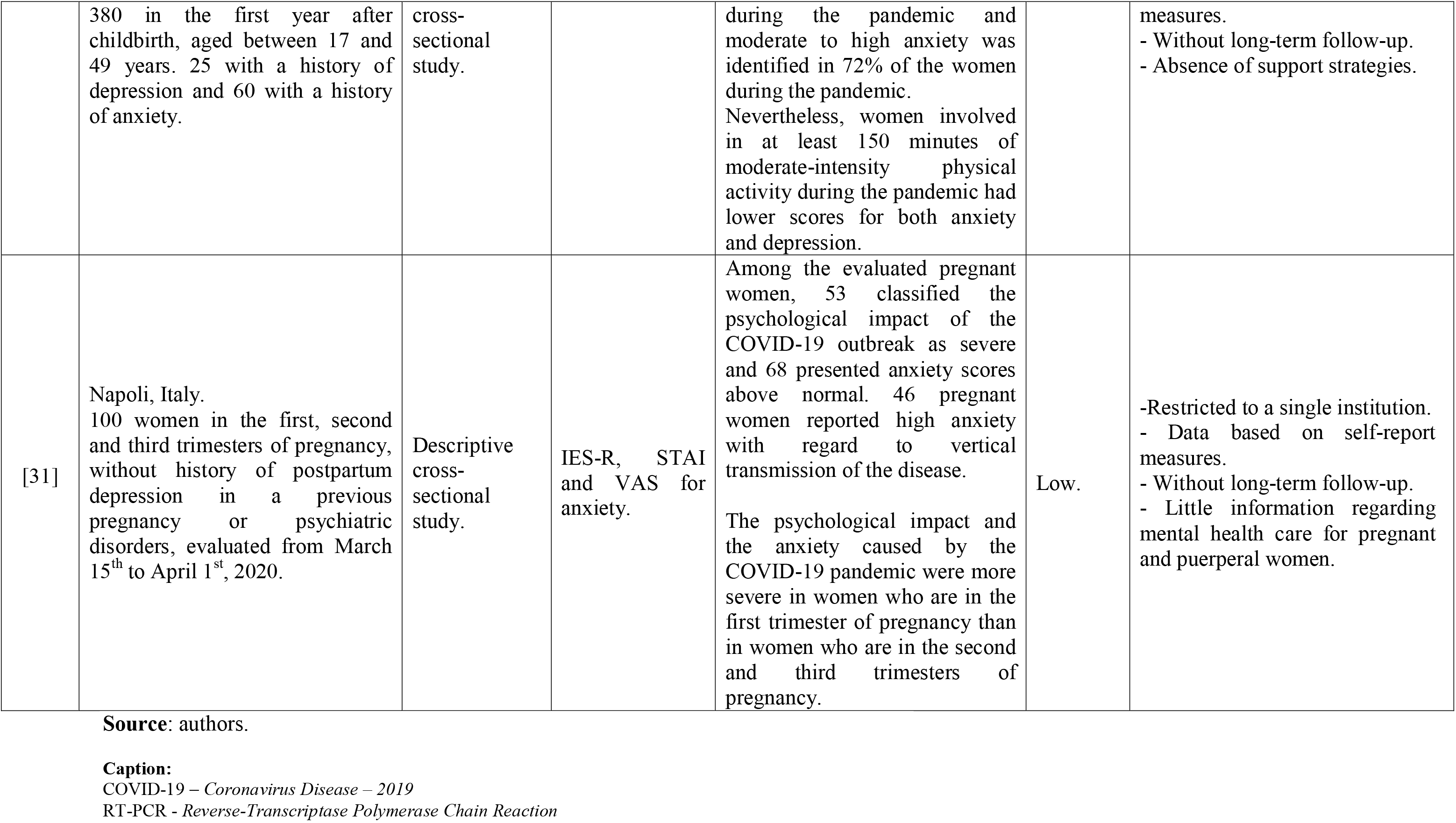
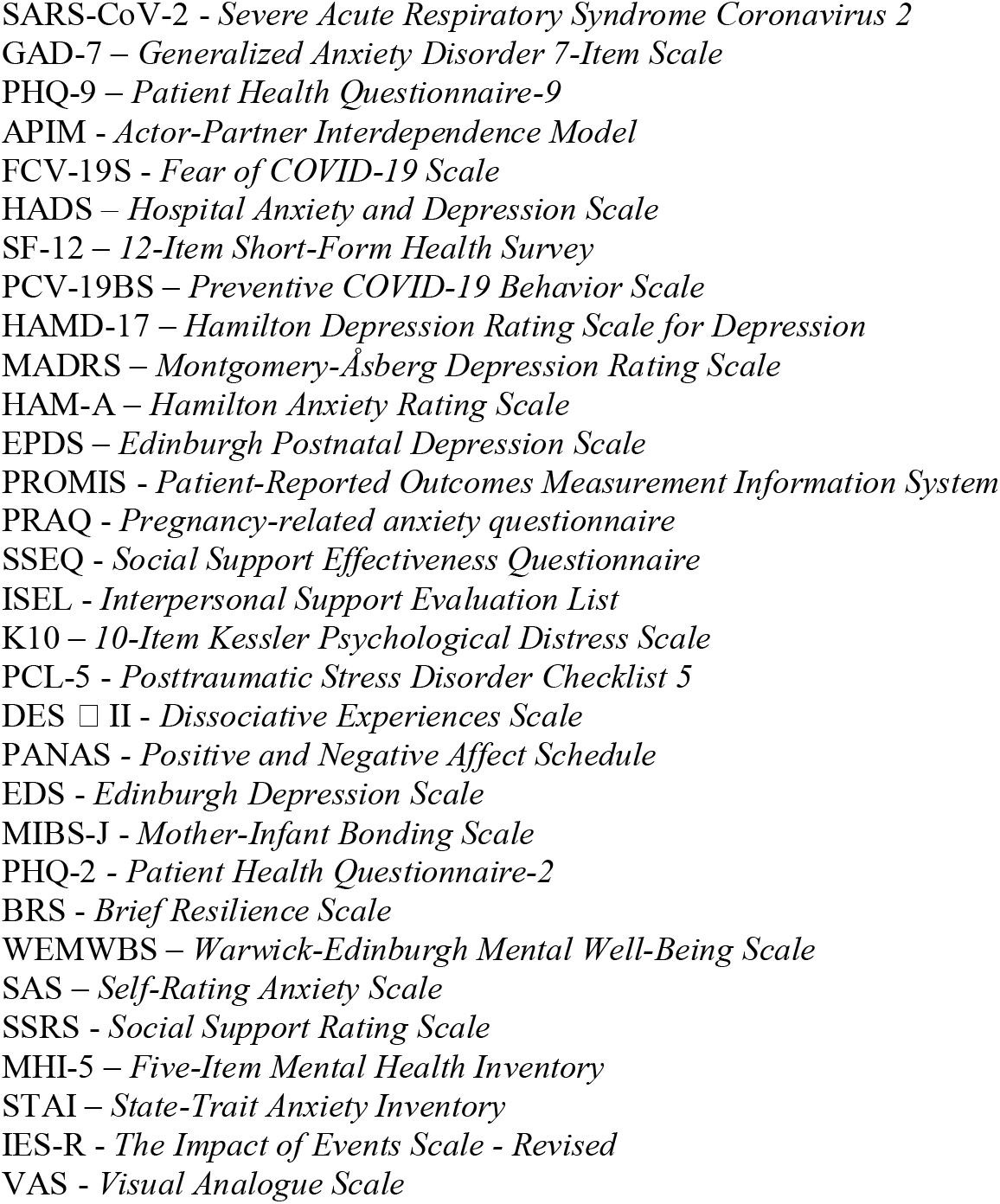
Characterization of the studies included in the final sample of this systematic review regarding the reference; the location and the population; the study design; the evaluation instrument; outcomes; the level of evidence and; the main limitations. Ribeirão Preto, SP, Brazil, 2020.

The analysis of the articles enabled us to observe the following characteristics: all articles were published in the year 2020, carried out in 13 different countries, including the United Kingdom [14,20], Iran [15], India [16], China [17,19,25], Canada [18,21,30], Belgium [22], Japan [23], United States [24], Italy [26,31], Australia [27], Ireland [28] and Israel [29].

A total of 10 studies adopted a descriptive cross-sectional design [14,15,18,20,22,25,27,28,29,31], one multicenter cross-sectional [19], one analytical cross-sectional [23], one analytical and multicenter cross-sectional [30], three case reports [16,17,26], one from a prospective cohort [21] and one from a mixed exploratory method [24].

Anxiety and depression were the main identified outcomes, being shown in 15 [14,16,17,18,19,20,21,22,24,25,27,28,29,30,31] and 11 studies [14, 15,17,18,19,20,21,22,23,24,30], respectively. Other outcomes identified in more than one study were: concerns related to several factors [17,18,27,28]; loneliness [18,20,24]; stress [24,27] and fear [15,16]. The studies evaluated anxiety from several scales: three studies using the *Generalized Anxiety Disorder 7-Item Scale* – GAD-7 [14,22,24], two using the *State-Trait Anxiety Inventory* – STAI) [30,31], one using the *Hospital Anxiety and Depression Scale* – HADS [15] and one using the *Hamilton Anxiety Rating Scale* – HAM-A) [17]. Another study also used the Self-Rating Anxiety Scale – SAS [25], and another the Pregnancy-related Anxiety Questionnaire – PRAQ [18]. Moreover, two studies evaluated anxiety using specific instruments [20,29].

Depression was evaluated in: one study using the *Patient Health Questionnaire-9* – PHQ-9 [14], one using the *Hospital Anxiety and Depression Scale* – HADS) [15], one using the *Hamilton Depression Rating Scale for Depression* – HAM-D-17 [17], one using the *Montgomery-Åsberg Depression Rating Scale* – MADRS [17], four studies using the *Edinburgh Postnatal Depression Scale* – EPDS [18,19,23,30], two using the *Patient Health Questionnaire-2* – PHQ-2 [17,24] and another study using the *Edinburgh Depression Scale* (EDS) [22]. Another study also used its own instrument to evaluate depression [20].

Moreover, the fear of COVID-19 was evaluated in one study using the *Fear of COVID-19 Scale* – FCV-19S) [15]; mental well-being in one study using the *Warwick-Edinburgh Mental Well-Being Scale* – WEMWBS) [24]; quality of life in one study using the *12-Item Short-Form Health Survey* – SF-12) [15]; preventive behavior in one study using the *Preventive COVID-19 Behaviour Scale* (PCV-19BS); psychological distress in one study using the *10-Item Kessler Psychological Distress Scale* – K10) [21]; posttraumatic stress disorder in two studies, one using the *Posttraumatic Stress Disorder Checklist 5* – PCL-5 [21] and the other using the *Impact of Events Scale–Revised* – IES-R) [31]; positive and negative affects in one study using the *Positive and Negative Affect Schedule* – PANAS) [21]; one study evaluated social support through the Social Skills, Behavior Problems and Academic Competence Inventory for Children (SSRS) [25] and two studies evaluated psychological stress and well-being through the *Five-Item Mental Health Inventory* – MHI-5 [29] and the *Visual Analogue Scale* – VAS [31], respectively.

Among the 18 analyzed studies, 11 signalized a low level of evidence [14,16,18,19,20,21,22,28,29,30,31] and seven very low levels [15,17,23,24,25,26,27].

## 4. DISCUSSION

The results showed that the study population was mentally affected by the onset of symptoms of anxiety, depression, fear, suicidal thoughts, psychomotor agitation, irrelevant speech, crying spells, delusions of persecution and greatness, catatonic symptoms of mutism, bad mood, irritability, excessive concerns, thoughts of self-mutilation, social isolation, loneliness, stress, resilience and poor well-being.

Depression, fear, anxiety and other psychotic symptoms referred to are configured in the main psychological repercussions that affect the mental health of pregnant and puerperal women during the pandemic period of COVID-19, thus reflecting the sociopolitical-cultural context in which we live and its challenging capacity. The current pandemic scenario triggered by COVID-19 and the consequent mitigation measures (social detachment, quarantine and social isolation) have entailed the intensification of psychological distress for certain groups, which can include women in the pregnancy-puerperal cycle.

This is because pregnancy and postpartum, commonly, are already characterized as periods of hormonal, bodily and psychological changes in women’s lives, adding, in the current scenario, a pandemic disease with transmission routes still unclear, but that can threaten maternal and child health. Thus, the experience of a continuous pandemic situation and the lack of certainty about the future at this moment can be factors causing stress, anxiety, fear and other symptoms, since without information, they do not know how to proceed to protect themselves and their babies. This situation has already been reported as recurrent in outbreaks related to other infectious diseases such as H1N1 Influenza, Ebola, *Middle East Respiratory Syndrome* (MERS), *Severe Acute Respiratory Syndrome* (SARS) and Zika Virus [32,33].

Although the vertical transmission of *Severe Acute Respiratory Syndrome Coronavirus 2* (SARS-CoV-2) is not clear, we cannot rule out: concerns about the possibility of premature birth; the chances of cesarean section in cases of idealized vaginal birth; the lack of humanization during childbirth; the impossibility of breastfeeding; the newborn being isolated from the mother, or the mother and the newborn being isolated from family members and close acquaintances; besides the fear of being infected or the baby being or being infected by other people [32,33].

The fear of being infected by COVID-19 is very distressing, mainly because it still does not have scientifically proven treatment. Social detachment, a primary measure to contain the proliferation of the virus, made people physically isolate themselves from family members, friends, their tasks and the community, a measure that affected the mental health and psychological well-being of pregnant and puerperal women [18].

In this sense, depressive symptoms were widely identified in studies [14,15,17,18,19,20,21,22], despite the limiting difference observed in culture studies, methods and instruments used, as well as in the fear of being infected by infectious diseases, which was significantly associated with depression, stress and anxiety [15,17,18,19,20,21,22, 27,28,29,30,31], where only one study performed in the United Kingdom [14] with women aged 18 to 39 years, from the 27^th^ to the 39^th^ weeks of pregnancy, presented a difference in the results, and this was due to the increase in information available about COVID-19 and to the guarantee offered to the population in question with respect to primary care and health professionals.

Symptoms of stress, depression and anxiety related to COVID-19 have shown a negative effect, thus impacting the mental health of the general population [34,35,36]. A research conducted at the beginning of the COVID-19 outbreak in Wuhan, China, found that 53.8% of the participants presented symptoms of anxiety and depression (moderate and severe), 17% and 29%, respectively [6].

Psychological disorders such as depression and anxiety, more present in the lives of pregnant and puerperal women in this pandemic period, have caused serious problems, with a consequent reduction in the quality of life of these individuals. Thus, there is a need to guarantee emotional support, in order to mitigate the fear of this illness [18]. Increased social and emotional support contributes to the reduction of psychological distress and to the maintenance of physical well-being, as well as family and partner support is of utmost importance in this process of social detachment [15], in addition to the issue of resilience [14], which helps us to cope well with severe situations such as the pandemic period.

Conversely, women without history of pre-existing mental illnesses who did not have support and suffered external pressure caused by third parties had psychotic breaks [16], others with a family history of bipolar disorder and, without support, also had psychotic characteristics [21,26]. Given this situation, we should observe that the infection and the fear of COVID-19 go beyond the organic issue, pointing to mental and social issues, by generating psychological exhaustion, especially for the most vulnerable people who feel unsupported.

One of the studies raised in this review pointed out that mental health problems are prevalent in women with younger ages, low family income and lower schooling level [21]. Common Mental Disorders (CMD) are challenging for health services, as they have grown alarmingly in the last few years. In particular, the occurrence of these disorders has increased in women, since they are twice as likely to develop the disorder as men, mainly when they are in the periods of pregnancy and puerperium [37].

Women in situations of risk of vulnerability, poverty, without a steady partner, young, with hereditary or congenital characteristics and low schooling levels have evolved with depressive and anxious symptoms not only in maternal health, but also fetal, which highlights the importance of screening and following-up during pregnancy and puerperium [38,39,40,41].

Moreover, many women experienced increased psychological stress, as they did not have the necessary help in the service unit, as well as support when breastfeeding. After childbirth, women may feel helpless, where the lack of support is a factor that can directly affect the breastfeeding process. [21].

Based on studies by Thomson, Ebisch-Burton and Flacking [42], it is common for women to have the need to carry out breastfeeding, so that those who do not can feel that they have failed. Nevertheless, according to the authors, both mothers who do not breastfeed and those who breastfeed may suffer from the absence of support and inconvenient judgment, which can even result in feelings of inadequacy and isolation [42]. Such notes show that the scenario related to COVID-19 may be the aggravating factor in the face of the issue that involves complications and the need for attention even outside the pandemic period.

Some limitations should be considered when interpreting the results of this systematic review. We noted a heterogeneity in the studies, the use of different instruments for data collection and distinct follow-up times.

## 5. CONCLUSION

From this review, we can infer that the COVID-19 pandemic has impacted the mental health of pregnant and puerperal women. The results have revealed the main changes in the mental health of this population, with depression and anxiety being the most frequent. Several concerns, loneliness, fear, stress, suicidal thoughts, psychosis, agitation, delusions of persecution and overvalued ideas were also present.

The current pandemic scenario triggered by COVID-19 and the consequent social detachment, the media pressure, the fear of contracting the infection, the economic scenario and the rupture of family rituals resulted in observable intensifications of psychological distress, thus causing changes in the mental health of these women.

## Data Availability

Available upon request

